# Dispersal history of SARS-CoV-2 variants Alpha, Delta, and Omicron (BA.1) in Spain

**DOI:** 10.1101/2024.07.01.24309632

**Authors:** Pilar Gallego-García, Samuel L. Hong, Nena Bollen, Simon Dellicour, Guy Baele, Marc A. Suchard, Philippe Lemey, David Posada

## Abstract

Different factors influence the spread of SARS-CoV-2, from the inherent transmission capabilities of the different variants to the control measurements put in place. Here we studied the introduction of the Alpha, Delta, and Omicron-BA.1 variants of concern (VOCs) into Spain. For this, we collected genomic data from the GISAID database and combined it with connectivity data from different countries with Spain to perform a phylodynamic Bayesian analysis of the introductions. Our findings reveal that the introductions of these VOCs predominantly originated from France, especially in the case of Alpha. As travel restrictions were eased during the Delta and Omicron-BA.1 waves, the number of introductions from distinct countries increased, with the United Kingdom and Germany becoming significant sources of the virus. The largest number of introductions detected corresponded to the Delta wave, which was associated with fewer restrictions and the summer period, when Spain receives a considerable number of tourists. This research underscores the importance of monitoring international travel patterns and implementing targeted public health measures to manage the spread of SARS-CoV-2.

## Introduction

The COVID-19 pandemic caused by SARS-CoV-2 resulted in governments worldwide imposing control measures of different severity. The extent of these restrictions varied with time and country and, in some cases, by region (Baniasad et al., 2021; Bergquist et al., 2020; European Union Agency for Fundamental Rights, 2020). Such was the case in Spain, with its decentralized healthcare system that allowed each of its 17 regions or autonomous communities to impose their own limitations. Thus, depending on the period and region, the COVID-19 measures in Spain were more or less restrictive (García-García et al., 2022). Apart from those regional measures, the Spanish central government also imposed several nationwide restrictions, such as a general lockdown and mandatory mask-wearing, and coordinated the vaccination campaigns in the different regions. SARS-CoV-2 sequencing efforts in Spain (Figure 1A) also varied with time and region, leading to regional differences in the number of sequences available for each period.

**Figure 1.**
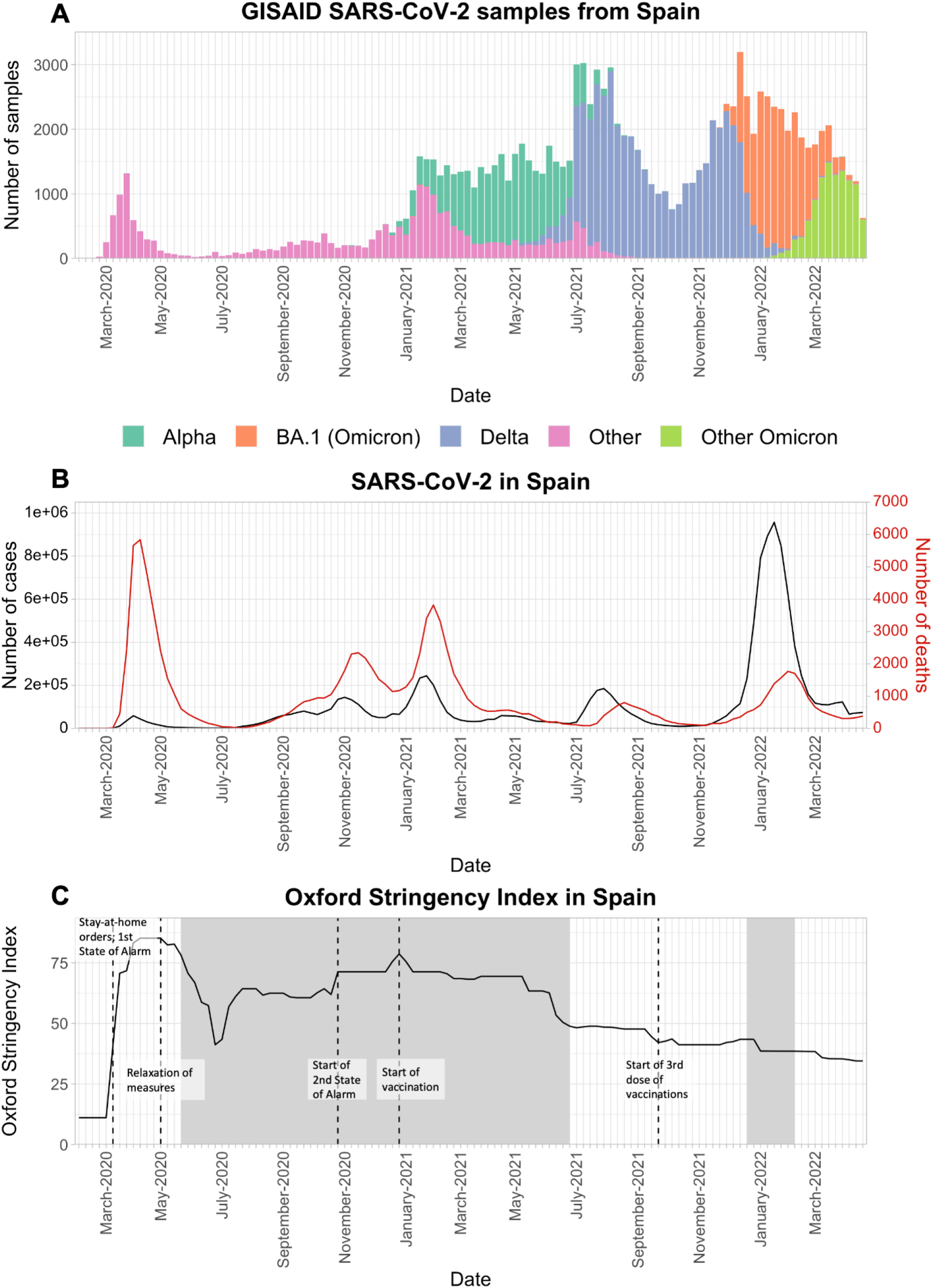
SARS-CoV-2 epidemic waves in Spain from February 2020 to April 2022. A) Number of sequences from Spain available in the GISAID database until June 11, 2022. Bar colors represent different lineages or variants, as explained in the legend. B) Number of diagnosed cases (left-axis; black line) and deaths (right-axis; red line) of COVID-19 in Spain. Data taken from OurWorldInData (Mathieu et al., 2020). C) Spain’s Oxford Stringency Index (Hale et al., 2021), representing how strict the measures were in Spain at any given moment (the index represents the response level of the strictest region within Spain). Data taken from OurWorldInData (https://ourworldindata.org/). Shaded areas represent periods in which masks were mandatory outside.

From the start of the pandemic up to the beginning of 2022, Spain suffered six epidemic waves of COVID-19 (Figure 1B). The first wave, which started around March 2020, following the first introductions of SARS-CoV-2 into the country, resulted in a national lockdown imposed to limit viral transmission in order to avoid overwhelming the healthcare system. Despite not being the most significant wave in terms of the number of cases, it was the largest in terms of deaths. Several factors contributed to these statistics, such as the novelty of the disease, for which there was no vaccine or previous immunity, an aging population, and the virus’ early entrance into nursing homes.

After the lifting of the lockdown on June 21, 2020, tied to the end of the first epidemic wave, the loosened restrictions (Figure 1C) favored the start of a new wave of infections towards the end of the summer (Figure 1B). This second wave was associated with the spread of the B.1.177 lineage from Spain and across Europe (Hodcroft et al., 2021; Lemey et al., 2021) and lasted until the end of 2020. Around this time, the Alpha variant of concern (VOC) was introduced in Spain, spreading with the rest of the preexisting variants and dominating the third wave. Despite the containment measures, increased social connections during the Christmas holidays likely sustained this spread, which resulted in the highest peak of cases until that point (Iftimie et al., 2022). The pre-existing immunity and the stringent containment measures put in place, probably aided in limiting the extension of the fourth wave towards April 2021, also dominated by Alpha (Figure 1A-B). While the vaccination campaign started around the same time as the first Alpha wave, it is not clear it was sufficiently advanced to have an effect on the posterior spread of Alpha. During the two Alpha epidemic waves, there were also some anecdotal detections of other VOCs (namely Beta and Gamma) or variants of interest (VOIs; e.g., Eta and Zeta) (Iftimie et al., 2022).

The fifth epidemic wave, linked to the introduction of the Delta VOC, occurred during the summer of 2021. It was potentially facilitated by the lifting of certain control measures, such as the outdoor mask-wearing mandates (Real Decreto-ley 13/2021, 2021), and resulted in a new peak in the number of cases, although lower than in the third wave. Subsequently, the end of 2021 encompassed the last moments of Delta and the start of the sixth wave, characterized by the introduction of a diversity of Omicron VOC lineages, starting with BA.1 and its sublineages (Figure 1A). This wave presented the highest number of detected cases of the pandemic in Spain and a significant number of deaths, although much less than in the first three epidemic waves.

Here, we focus on the study of waves three to six of the pandemic in Spain, considering the predominant VOCs during those periods: Alpha, Delta, and Omicron-BA.1. The first year of the pandemic in Spain has already been studied (García-García et al., 2022; Hodcroft et al., 2021; Iftimie et al., 2022; López et al., 2021), but the origin and importance of virus importations after 2020 have yet to be described. Hence, we here shed light on the events of the following waves (until the start of Spring 2022) that we subsequently discussed in the context of the control measures in place during those periods.

## Methods

### Subsampling

To study the history of Alpha, Delta, and Omicron-BA.1 introductions in Spain, we selected 7,699 worldwide SARS-CoV-2 whole-genome sequences from GISAID based on the prior probability of their sampling location resulting in introductions to Spain. To calculate this probability, we used aggregated Facebook mobility data (Maas, 2019) from January 2021 to March 2022 (Supplementary Figure S1), which reflects the number of Facebook users transiting from one country to another.

First, we computed the connectivity from all available countries in the Facebook data to Spain for each of the three studied periods (i.e. Alpha, Delta, and Omicron-BA.1) and ranked them from most to least connected to Spain. Then, we selected those most-connected countries that encompassed 95% of the total mobility to Spain. For each country, we sampled the number of sequences proportionately to the cumulative number of cases times the fraction of connectivity to Spain. We set an arbitrary threshold of 6.5 genomes per 10,000 cases, with a minimum number of sequences per country of 100 (see Truong Nguyen et al., 2022). For Omicron-BA.1, we set a maximum number of 500 sequences for each location for the analysis to remain computationally feasible, as the peak of cases in most countries increased the number of selected sequences.

We also included Delta sequences from India to account for the geographical origin of the Delta VOC (Vaidyanathan, 2021). We already had Alpha sequences from the United Kingdom (UK) (Kraemer et al., 2021) in our dataset, and for Omicron-BA.1, although the first cases were detected in South Africa, there is no clear country of origin (WHO, 2023). We could not obtain mobility data from India to Spain, so to compute the number of Indian sequences sampled, we used public information about the number of flights between Spain and India during the Delta period (Strohmeier et al., 2021).

In addition, we sampled 1,000 sequences from multiple Spanish regions, with the number of sequences per region chosen proportionately to the number of detected positive cases. As in previous studies (Truong Nguyen et al., 2022), we set these thresholds arbitrarily to obtain a manageable dataset that could be studied under a Bayesian framework within a reasonable computational time.

### Subsampling of SARS-CoV-2 sequences from GISAID

We downloaded the metadata for all the Alpha, Delta and Omicron-BA.1 sequences available in GISAID (Elbe & Buckland-Merrett, 2017) that met the following GISAID criteria: were “complete” (i.e., >29,000 nt); came from a human host; were associated with a complete collection date (year-month-day); and, for Alpha and Delta, were “high coverage” (<1% of missing bases and <0.05% of unique amino acid mutations). For Omicron-BA.1, we only excluded those with “low coverage” (>5% missing bases). However, we did not enforce the “high coverage” filter due to the substantial number of sequences that would have been discarded. We downloaded the Alpha and Omicron-BA.1 data on 2022-06-11 and the Delta data on 2022-04-26. The sampled periods were between weeks 52 of 2020 and 32 of 2021 (2020-12-20 to 2021-08-14) for Alpha, between weeks 25 and 50 of 2021 (2021-06-20 to 2021-12-18) for Delta, and between weeks 49 of 2021 and 13 of 2022 (2021-12-05 to 2022-04-03) for Omicron-BA.1.

Based on the metadata and the number of sequences determined from each country or Spanish region, we downloaded an even number of weekly sequences from GISAID. We only considered sequences that passed Nextclade quality tests and presented a “good” overall status, and which were assigned to the corresponding Nextclade clade and PANGO lineage (clade 20I / lineage B.1.1.7 for Alpha, clades 21A, 21I & 21J / lineage B.1.617.2 and sublineages (AY) for Delta, and clade 21K, lineage BA.1 and sublineages for Omicron). For Omicron, we limited the studied lineages to BA.1 and descendants because their pandemic wave was still ongoing when we initiated this study.

We used seqkit v2.1.0 (Shen et al., 2016) to remove all duplicates, aligned the sequences with Nextalign v.1.11.0 (Hadfield et al., 2018), and estimated a maximum likelihood phylogenetic tree with IQ-TREE v.2.1.3 (Minh et al., 2020), using 2,000 ultrafast bootstraps (Hoang et al., 2018) under the best-fit substitution model selected by ModelFinder (Kalyaanamoorthy et al., 2017). We then used TreeTime v.0.8.1 (Sagulenko et al., 2018) with the option “clock” to detect and subsequently remove “temporal outliers,” which involves sequences with divergences that are unexpected given their sampling time. We repeated this subsampling when possible until all sequences passed the filters. The final number of analyzed sequences was 3,024, 3,390, and 4,271 for Alpha, Delta, and Omicron-BA.1, respectively.

### Bayesian analysis of introductions

We inferred viral introductions in Spain using BEAST v1.10.5 (prerelease #23570d1) (Suchard et al., 2018). For these analyses, we followed the same strategy as Gallego-García et al. (2024) and employed a phylogeographic generalized linear model (GLM) in which we incorporated the country of origin of the sequences as a discrete trait and the connectivity matrix between those countries as a covariate (Gill et al., 2016; Lemey et al., 2021). The connectivity matrix was computed based on the Facebook international mobility data previously described and was log-transformed and standardized before being included in the analysis.

We performed Markov chain Monte Carlo (MCMC) analyses in BEAST using a timed tree estimated by TreeTime (with options --max-iter 10 and --reroot least-squares) as the starting tree, HKY as the substitution model (Hasegawa et al., 1985) with empirical base frequencies and rate variation among sites following a discretized gamma distribution with four rate categories (Yang, 1994). We fixed the evolutionary rate at 7.5 × 10^−4^ nucleotide substitutions/site/year (du Plessis et al., 2021) and used a Bayesian non-parametric coalescent (skygrid) model as tree prior (Gill et al., 2013). Missing covariate values in the connectivity matrices were integrated out using efficient gradient approximations (Didier et al., 2024) to the trait data likelihood (Ji et al., 2020). All the BEAST analyses were run in the Galician Supercomputing Center (CESGA) in combination with the BEAGLE (v.4) (Ayres et al., 2019) high-performance computational library to accelerate the computations.

We simulated a minimum of seven independent MCMC chains per dataset, discarded at least 10 percent of each chain as burn-in, and combined the results with LogCombiner (Suchard et al., 2018). We ran the chains until all continuous parameters presented effective sample sizes (ESSs) > 200 for Alpha and ESSs > 100 for Delta and Omicron-BA.1. After combining the chains, they totaled 638M generations for Alpha, 794M for Delta, and 1,388M for Omicron-BA.1. The transition histories and their timings were summarized from the posterior tree distributions using the tool TreeMarkovJumpHistoryAnalyzer provided within the BEAST codebase (Lemey et al., 2021; Minin & Suchard, 2008).

## Results

### Alpha

The analyses revealed that during the Alpha wave period, there were at least 99 independent introduction events (95% highest posterior density [HPD]: 83-113) from other countries to Spain (Figure 2A-B). By far the most introductions to Spain during the Alpha period came from France (83, 95% HPD: 70-97). However, most introductions produced fewer than ten sampled descendants (84, 95% HPD: 68-96). Of those, around half were singletons, meaning there was only one sampled descendant (48, 95% HPD: 39-56), and thus most introductions did not produce substantial local transmission. On the other hand, 15 introductions (95% HPD: 11-19) resulted in a sizeable number of descendants (i.e. > 10 sampled tip nodes) and these introductions originated almost exclusively in France.

**Figure 2.**
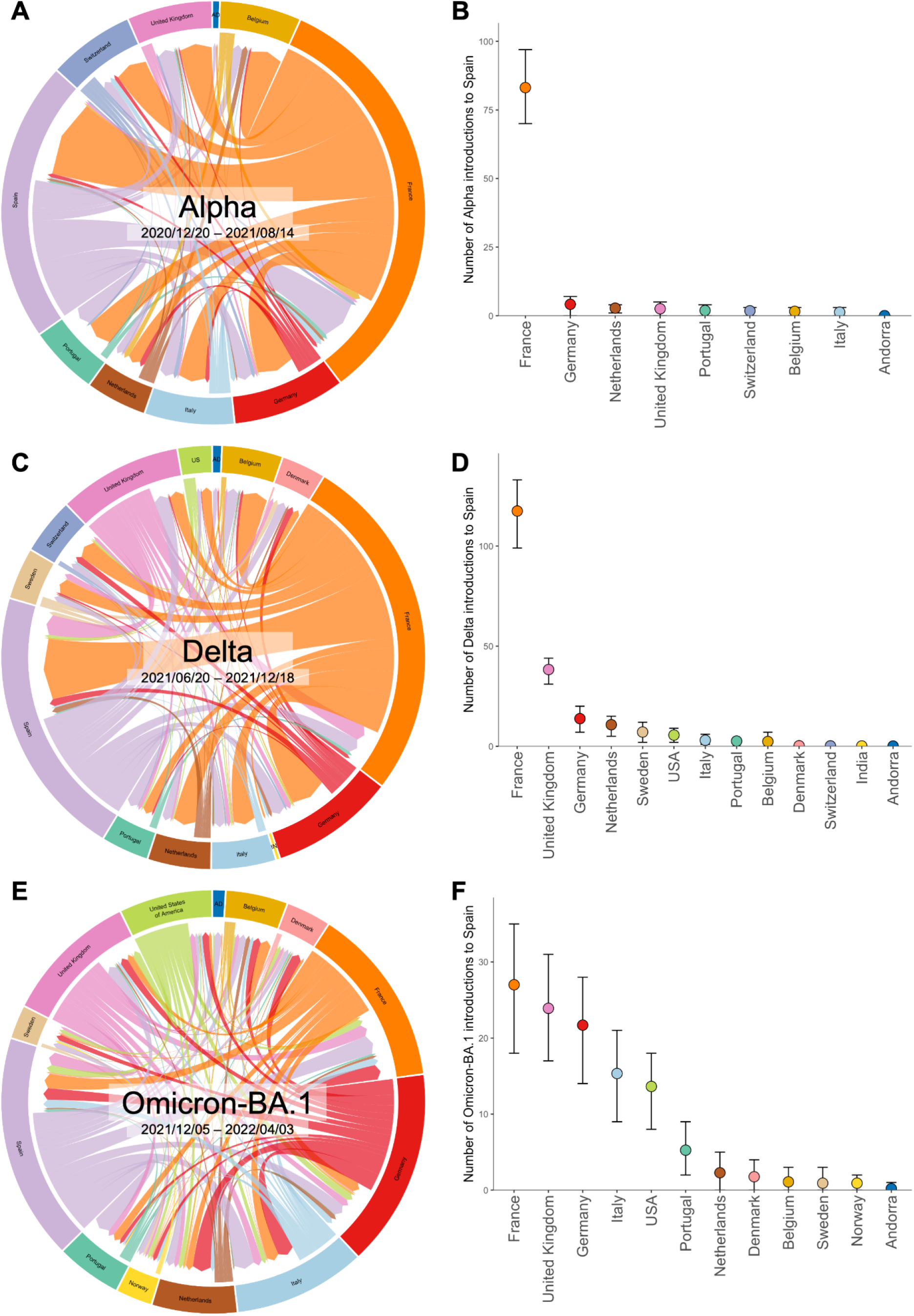
SARS-CoV-2 introduction events into Spain. Circular migration flow plots based on the Markov jumps between different countries and Spain (A: Alpha; C: Delta; E: Omicron-BA.1). Mean and 95% highest posterior density (HPD) number of transitions from those countries to Spain, computed from all the trees from the posterior distribution (B: Alpha; D: Delta; F: Omicron-BA.1). Codes ISO 3166-2 AD, IN, and US correspond with Andorra, India, and the USA respectively.

### Delta

In comparison with Alpha, during the Delta period there was an increase in the number of introductions and the number of originating countries (Figure 2C-D). We detected 202 (95% HPD: 186-221) introductions, with France again being the most important source of viral introductions (117, 95% HPD: 99-133). The number of introductions from the UK increased significantly (38, 95% HPD: 31-44), followed by Germany (14, 95% HPD: 7-20) and the Netherlands (11, 95% HPD: 5-15). There were also introductions from the USA, Italy, Portugal, and Belgium. Again, most introductions did not lead to substantial transmission clusters (187 tip nodes, 95% HPD: 170-205), with most being singletons (127, 95% HPD: 112-142). However, we did not detect introductions from India to Spain (or vice versa), despite it being the geographical origin of the Delta variant (Figure 3B).

### Omicron-BA.1

In the case of the first wave of Omicron (BA.1 and its sublineages) (Figure 2E-F), there was a decrease in the number of identified introduction events (114, 95% HPD: 100-128) compared to the Delta period, although in both periods many introductions were small (104, 95% HPD: 90-118), primarily singletons (76, 95% HPD: 62-88). France was again the primary source of introductions, with around 27 (95% HPD: 18-35) estimated introductions, but the number of detected introductions from the UK and Germany was quite similar, with 24 (95% HPD: 17-31) and 22 (95% HPD: 14-28) respectively. Italy and the USA were also important sources of Omicron-BA.1 introductions to Spain. In this case, we also detected at least five introductions (95% HPD: 2-9) of SARS-CoV-2 from Portugal to Spain.

## Discussion

The introductions of SARS-CoV-2 to Spain for the Alpha, Delta, and Omicron-BA.1 VOCs (epidemic waves three to six) reflect the changing landscape of COVID-19 restrictions and containment measures (Figure 1C), as well as the mobility peaks linked to vacation periods, notably around August (Supplementary Figure S1).

Contrary to the previous SARS-CoV-2 waves, most of the Alpha, Delta, and Omicron-BA.1 introductions came from France. For example, for the first wave, most of the detected introductions came from Italy, the Netherlands, England, and Austria (López et al., 2021). This large contribution from France could have been shaped by the combination of a high number of cases and a high connectivity to Spain, making it the country with the largest number of representative sequences in this study, after Spain. However, we also have to take into account that, together with Portugal, which is much smaller and is connected primarily by terrestrial ways to Spain, France was the primary way of entrance to Spain from the rest of Europe, especially in a context where air connections were much more restricted. Peninsular Spain also shares borders with Andorra and the UK (via Gibraltar), but their much smaller population and territory renders their importance at a national level negligible.

This predominance of French introductions was especially noticeable during the Alpha period (third and fourth waves), when, despite the movement restrictions put in place by Spain and France, almost all of the introductions into Spain (>80%) came from France. At the same time, we detected markedly fewer introductions from other countries with high connectivity with Spain. Towards the Alpha period, Spain and France were under stringent containment measures, including curfews and perimeter closures that restricted mobility around their territories. Apart from this, movements across the border were restricted to essential ones, such as cross-border workers and freight transports, or required the presentation of a negative PCR test (or a vaccination certificate, when these became more widely available). Despite these aspects and the recommendations to reduce traveling, the Spanish-French border was not outright closed, so there could still be some mobility, allowing a steady exchange of travelers and, therefore, of viruses. The influx of viruses from France was especially prominent towards the beginning of the Alpha period and declined as time passed, without peaks that could be linked to holiday periods such as the Holy Week. On the other hand, the border with Portugal was closed from the end of January to the beginning of May, except for exceptional reasons, which could help explain why we did not detect almost any introductions from Portugal to Spain. We must also consider that Portugal is much less populated than Spain and France, which also relates to fewer infected people, and its geographical location, not bordering any other country. All these factors combined could help to explain Portugal’s limited role as a source of introductions, at least towards Spain.

Additionally, in the case of the UK, there were several extra restrictions from both sides to travel between Spain and the UK due to the latter being the origin of the Alpha VOC (Kraemer et al., 2021; Orden PCM/1237/2020, 2020). While these measures aimed to limit SARS-CoV-2 transmission and the number of introductions, they differed for both countries. While Spain banned UK travelers from entering the country unless they were Spanish nationals or residents, the UK imposed a self-quarantine period at return, in both cases combined with other restrictions in the national territory. According to our analyses, these measures could have helped limit direct introductions from the UK to Spain.

The peak of Alpha introductions took place towards the beginning of the study period, while during the last weeks, i.e. in summer, as mobility increased (Supplementary Figure S1A) and restrictions eased, it was already being replaced by Delta. Meanwhile, the sharp increase in introductions to Spain during the Delta wave could be linked to the fact that it started during the summer. This period was characterized by increased international tourism (Supplementary Figure S1B) and higher national mobility (Supplementary Figure S2), while the level of restrictions was the lowest until that point. Lifting the outdoor mask mandates and appropriate social distancing, combined with the relaxation derived from the vaccination rollout, likely helped to propel this peak in introductions as more people decided to travel. Also, at the end of September, towards the middle of the Delta period, with around 80% of the Spanish population already vaccinated (Supplementary Figure S3), restrictions were further relaxed, and several regions entered the so-called “new normality” without specific restrictions.

The increase in tourism and lowered restrictions is likely reflected in the fact that, although France still ranked first in the number of introductions, it only represented around 58% of all introductions, a sharp decrease compared to the Alpha period. On the other hand, lifting the travel restrictions from and to the UK could explain its second position as a virus source. The sources of introductions during this period were much more diverse than during the Alpha wave, which could also be related to the lower level of restrictions across Europe. The increased transmissibility of Delta (Campbell et al., 2021; Earnest et al., 2022), as well as the higher risk of infection by this variant after vaccination (Andeweg et al., 2023), could also help explain the rapid variant replacement. Despite this, the peak in cases was slightly lower for Delta, the number of deaths was dramatically reduced, and an even higher percentage of the introductions were non-expansive, which means that pre-existing immunity in the population (by previous infections or vaccination), as well as some of the measures in place, must have played a role in restricting Delta spread.

Lastly, the Omicron-BA.1 period or sixth wave, which was associated with lower national mobility than for the Delta period, albeit higher than for Alpha (Supplementary Figure S2) and not linked either with a significant peak in international tourism (Supplementary Figure S1C), showed fewer introductions than the Delta wave. Nonetheless, with the lowest stringency of measures in place, despite the mandatory use of the mask outdoors again, the number of detected introductions was still higher than for Alpha. The fact that most of the introductions were non-expansive appears to contrast with the number of cases of this wave, which was the highest of the whole pandemic in Spain. This higher number of cases, on the other hand, might be explained by the increased transmissibility of Omicron-BA.1 compared to the previous VOCs (Elliott et al., 2022), to a reduced sense of risk from the population, when everyone who wanted had already been vaccinated, and to the so-called “pandemic fatigue” (Du et al., 2022; World Health Organization. Regional Office for Europe, 2020).

The reasons for the limited number of Omicron-BA.1 introductions in Spain could be multiple. One possible explanation is that the Omicron period, taking only BA.1 into account, is significantly shorter than the Alpha and Delta period (35, 26, and 16 weeks for Alpha, Delta, and Omicron-BA.1, respectively). Additionally, the proportion of sequenced cases respective to the total cases was the lowest of the three waves (Supplementary Figure S4), as the sequencing reached peaks similar to Delta while the number of cases skyrocketed. This low proportion of sequenced cases could potentially limit the detection of introductions in the country, as many transmission chains could be undetected or underrepresented.

The sources of Omicron-BA.1 introductions were considerably more diverse than for previous VOCs. Although still in first place, France was only accountable for 24% of the introductions, and a very similar number of introductions was detected from the UK and Germany, in line with a lower level of preventive measures and travel limitations. We should note that even though the connectivity from France to Spain was much higher than that of the rest of the countries, the same number of Omicron-BA.1 sequences from France, Germany, Italy, and the UK were present in the study, as they reached the limit of 500 sequences per country of origin (Supplementary Table S1). The much larger number of initially selected sequences for this variant is related to the peak of cases that Omicron caused in the connected countries, as well as in Spain, and limiting those numbers was necessary to maintain the analysis computationally feasible. Even so, we expect the number of sequences from each country in the study to have a limited influence over the number of introductions as, for example, for Delta we included more UK sequences than French ones. Still, we detected three times as many introductions from France as from the UK. We should then consider that the proportion of German and British travelers that caused an introduction of SARS-CoV-2 in Spain had to be significantly higher and could be related to the different anti-COVID-19 measures put in place by each region, as the regions that receive most of those travelers are not the same. In the case of France, the mobility was higher towards Catalonia (Supplementary Figure S5A), in the northeast of Spain and bordering France, which imposed strict control measures during the Christmas period and January, including nocturnal curfews, the closure of nightlife and the mandatory presentation of the COVID-19 certificate. On the other hand, the highest mobility from Germany and the UK occurred towards the Canary Islands (Supplementary Figure S5B-C), which only required presenting the COVID-19 certificate. These differences in mobility trends, coupled with the asymmetric containment measures within Spain, could explain the lower proportion of French introductions compared to the previous waves and possibly highlight the importance and impact of those regional measures.

One trend common to the three VOCs studied was that most introductions produced only a handful of descendants. This could be due to the various prevention and containment measures, which greatly limited the interaction between people without masks for long periods, reducing the odds of a massive transmission event. Apart from this, quarantines after testing positive were mandatory at the beginning and then at least recommended, as well as maintaining social distancing and mask-wearing, which could also help deter transmission. This situation is also typical of SARS-CoV-2 transmission, as there is significant variability in the number of descendants produced, which means most of the introductions into a territory tend to make a low number of descendants, while some others are responsible for most of the cases (Borges et al., 2022; Dellicour et al., 2023; Tsui et al., 2023). It is also important to highlight that most of the detected introductions happened in the first stages of the studied periods or even a bit before them, so these variants may have already been circulating in the country before they were detected.

The number of introductions into Spain per variant also highlights the different impacts of external introductions during the pandemic, depending on the country and the specific VOC. In the case of Delta, having detected 202 individual introductions after using 1,000 Spanish sequences, it nearly doubles the capacity of Alpha and Omicron-BA.1 of establishing local transmission chains and is significantly higher than the one observed in the UK for the same variant (1,458 detected introductions for 52,992 sampled sequences; (McCrone et al., 2022). Conversely, the contribution of the Omicron-BA.1 introductions was only slightly higher relative to the UK (6,455 detected introductions for 81,039 sampled sequences; (Tsui et al., 2023) but lower than in Mexico (160 detected introductions for 641 sequences; (Castelán-Sánchez et al., 2022).

Lastly, we acknowledge that the mobility data we used present limitations as they depend on the number of users sharing their location with the application and, therefore, may only be representative of some of the population. Nonetheless, this type of data has already been used to study SARS-CoV-2 transmission linked to human mobility (Kraemer et al., 2020; Lemey et al., 2021; Truong Nguyen et al., 2022), and can still be considered a good approximation that should reduce the sampling bias in our analysis. Indeed, other types of sampling bias still exist, such as the difference in sequencing effort and strategy between countries and between Spanish regions, which can be substantial (Supplementary Figure S6). Despite this, in most cases, we believe the number of sequences available relative to the number of sequences used was large enough to ensure an adequate distribution of sampling dates and locations, mitigating this source of bias.

To conclude, the analysis of the SARS-CoV-2 introductions in Spain suggests that the control measures significantly reduced further importations of the virus into the country. Moreover, according to our analysis, the number of cases does not correlate with the number of introductions, and the relatively small number of descendants detected for most introductions suggests a limited impact on the progression of the pandemic.

## Data Availability

All the genomic sequences and associated metadata used in this study have been published in GISAID EpiCoV database (https://gisaid.org/) (EPI_SET_ID: EPI_SET_240610da, Supplementary Table S2).

https://gisaid.org

## Acknowledgments

We gratefully acknowledge all data contributors, i.e., the authors and their originating laboratories responsible for obtaining the specimens and their Submitting laboratories for generating the genetic sequence and metadata and sharing via the GISAID Initiative. PGG was supported by grant ED481A-2021/345 from the Consellería de Cultura, Educación e Universidade Xunta de Galicia. SD acknowledges support from the *Fonds National de la Recherche Scientifique* (F.R.S.-FNRS, Belgium; grant no. F.4515.22). SD and GB acknowledge support from the Research Foundation – Flanders (*Fonds voor Wetenschappelijk Onderzoek – Vlaanderen*, FWO, Belgium; grant no. G098321N) and from the European Union Horizon RIA 2023 project LEAPS (grant no. 101094685). GB acknowledges support from the Internal Funds KU Leuven (Grant No. C14/18/094), from the Research Foundation – Flanders (*Fonds voor Wetenschappelijk Onderzoek – Vlaanderen*, FWO, Belgium; grant no. G0E1420N) and from the DURABLE EU4Health project 02/2023-01/2027, which is co-funded by the European Union (call EU4H-2021-PJ4; grant no. 101102733). SD and PL acknowledge support from the European Union Horizon 2020 project MOOD (grant agreement no. 874850). PL and MAS acknowledge support from the European Union’s Horizon 2020 research and innovation programme (grant agreement no. 725422 – ReservoirDOCS), from the Wellcome Trust through project 206298/Z/17/Z and from the National Institutes of Health grants R01 AI153044, R01 AI162611 and U19 AI135995. PL also acknowledges support from the Research Foundation – Flanders (*Fonds voor Wetenschappelijk Onderzoek – Vlaandere*n, G0D5117N, and G051322N).

## Declaration of interest

There are no conflicting interests.

## Data availability

All the genomic sequences and associated metadata used in this study have been published in GISAID’s EpiCoV database (https://gisaid.org/) (EPI_SET_ID: EPI_SET_240610da, Supplementary Table S2).

## Notes

### Competing Interest Statement

The authors have declared no competing interest.

