## Supplemental Data for "Dispersal history of SARS-CoV-2 variants Alpha, Delta, and Omicron (BA.1) in Spain"

[1] CINBIO, Universidade de Vigo, 36310 Vigo, Spain
[2] Galicia Sur Health Research Institute (IIS Galicia Sur), SERGAS-UVIGO
[3] Department of Microbiology, Immunology and Transplantation, Rega Institute, Laboratory of Clinical and Epidemiological Virology, KU Leuven – University of Leuven, 3000 Leuven, Belgium
[4] Spatial Epidemiology Lab (SpELL), Université Libre de Bruxelles, 1050 Brussels, Belgium
[5] Department of Biostatistics, Fielding School of Public Health, University of California Los Angeles, Los Angeles, CA 90095, USA
[6] Department of Human Genetics, David Geffen School of Medicine, University of California Los Angeles, Los Angeles, CA 90095, USA
[7] Department of Computational Medicine, David Geffen School of Medicine, University of California Los Angeles, Los Angeles, CA 90095, USA
[8] Global Virus Network (GVN), Baltimore, MD 21201, USA
[9] Department of Biochemistry, Genetics, and Immunology, Universidade de Vigo, Vigo 36310, Spain

### Supplementary data

**Table S1.** Number of sequences selected before applying restrictions and finally used per country for the Omicron-BA.1 variant. We set a limit of 500 sequences and a minimum of 100 per country. For Andorra, we used all the sequences available in GISAID that passed the quality filters at that moment.

| Country | No. of sequences before restrictions | Final no. of sequences |
|---|---|---|
| France | 3550 | 500 |
| UK | 1379 | 500 |
| Portugal | 234 | 234 |
| Italy | 522 | 500 |
| Germany | 655 | 500 |
| Netherlands | 158 | 158 |
| Belgium | 52 | 100 |
| Andorra | 2 | 24 |
| Sweden | 22 | 100 |
| USA | 458 | 458 |
| Denmark | 38 | 100 |
| Norway | 13 | 100 |

**Table S2. GISAID accession data.**

Data Availability

GISAID Identifier: EPI_SET_240610da

doi: 10.55876/gis8.240610da

All genome sequences and associated metadata in this dataset are published in GISAID's EpiCoV database. To view the contributors of each individual sequence with details such as accession number, Virus name, Collection date, Originating Lab and Submitting Lab, and the list of Authors, visit 10.55876/gis8.240610da

Data Snapshot

- EPI_SET_240610da is composed of 10,685 individual genome sequences.
- The collection dates range from 2020-12-21 to 2022-04-03;
- Data were collected in 16 countries and territories;
- All sequences in this dataset are compared relative to hCoV-19/Wuhan/WIV04/2019 (WIV04), the official reference sequence employed by GISAID (EPI_ISL_402124). Learn more at https://gisaid.org/WIV04.

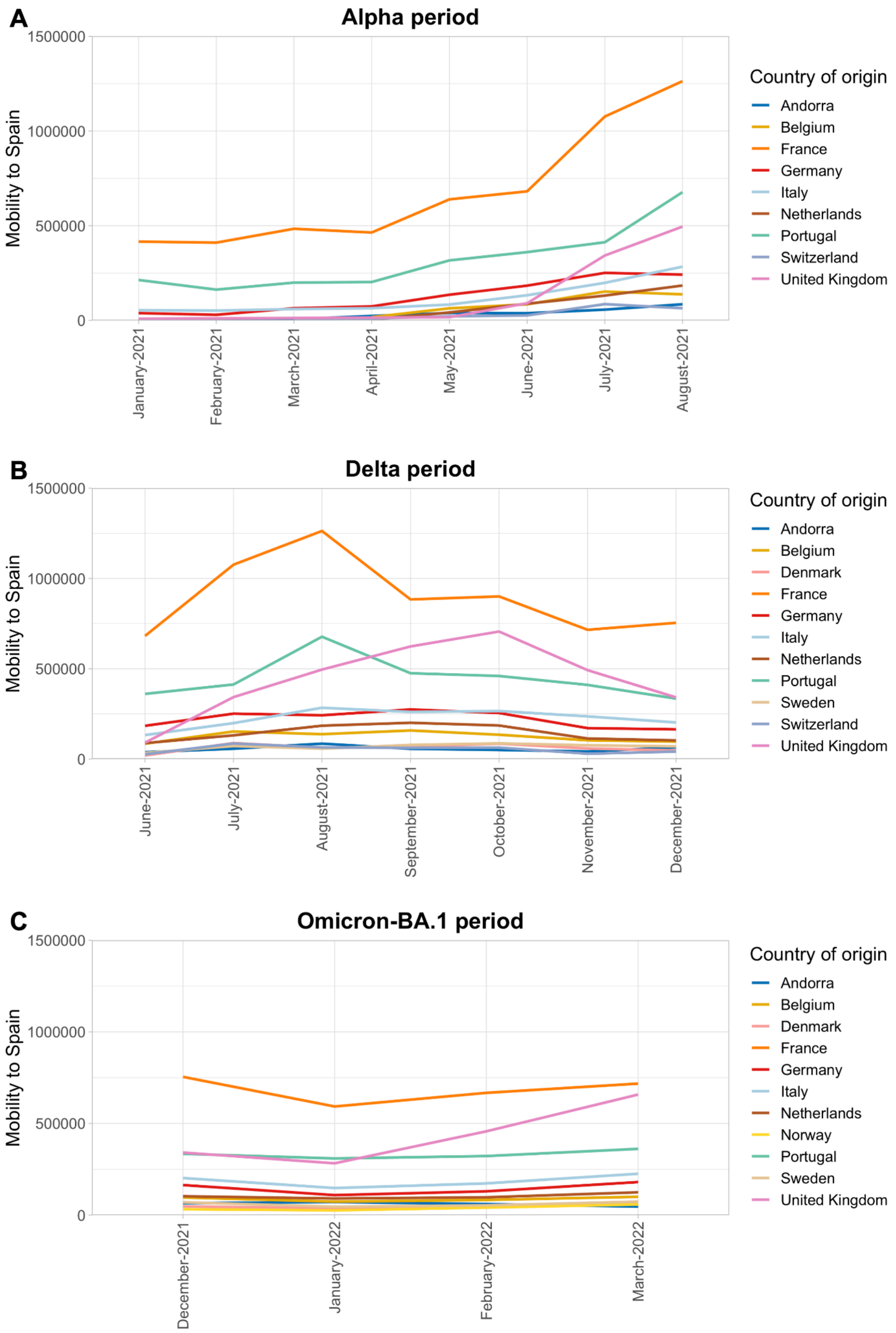


**Figure S1. Mobility to Spain from each studied country**: A) Alpha period; B) Delta period; C) Omicron-BA.1 period. Data were taken from the aggregated Facebook international mobility dataset.

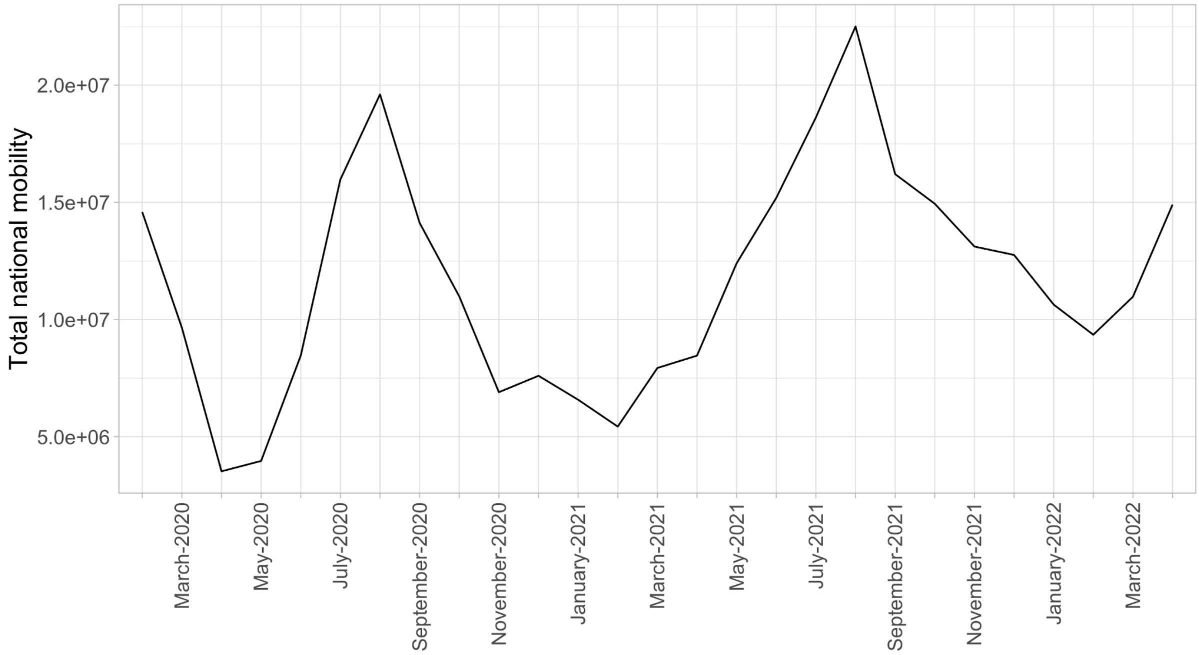


**Figure S2. Total national mobility in Spain from February 2020 to April 2022.** The original data were taken from the Spanish National Institute of Statistics (INE; https://www.ine.es).

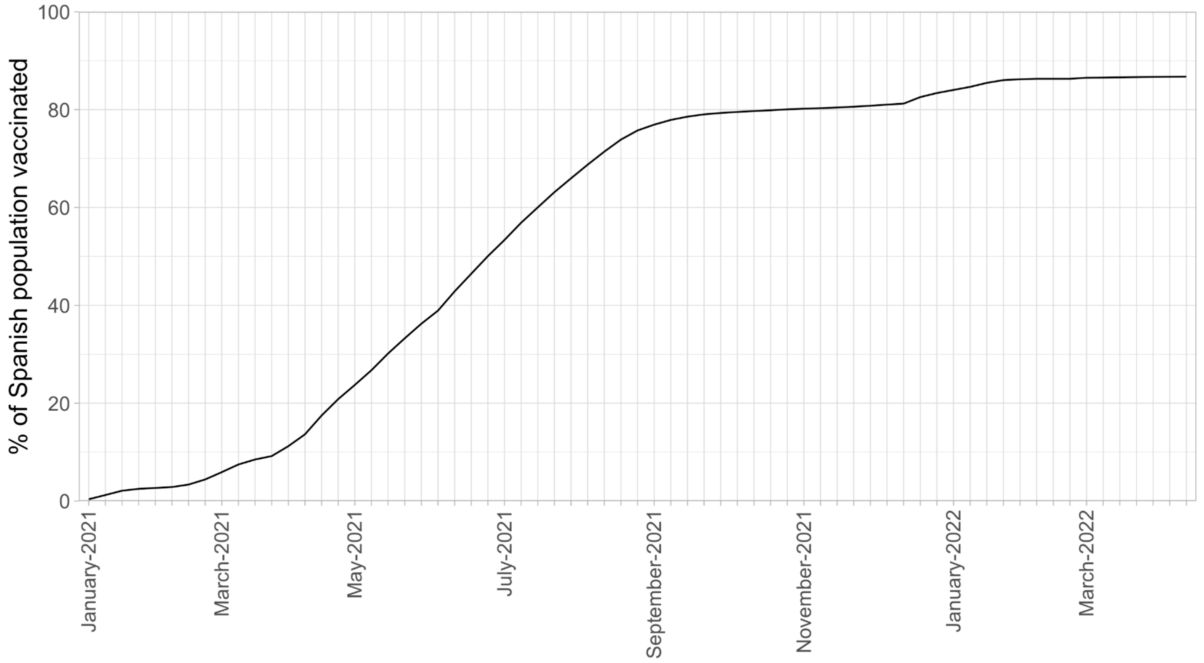


**Figure S3. Percentage of people in Spain who had received a complete vaccination schedule** (two doses for vaccines Moderna, Oxford/AstraZeneca, Pfizer/BioNTech and one dose for Johnson&Johnson/Janssen). Data taken from OurWorldInData (Mathieu et al., 2021).

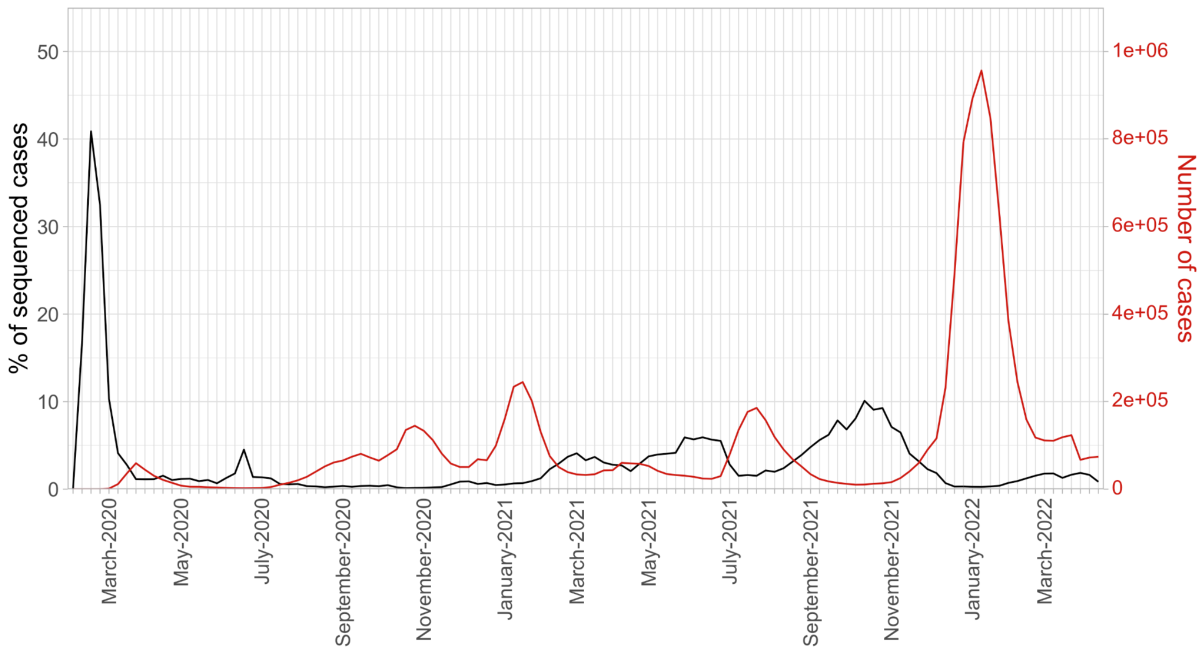


**Figure S4. Proportion of sequenced cases vs. total cases by date.** Data regarding total cases were taken from OurWorldInData (Mathieu et al., 2020). The percentage of sequenced cases was computed considering the number of sequences from Spain available in GISAID until 2022-06-11.

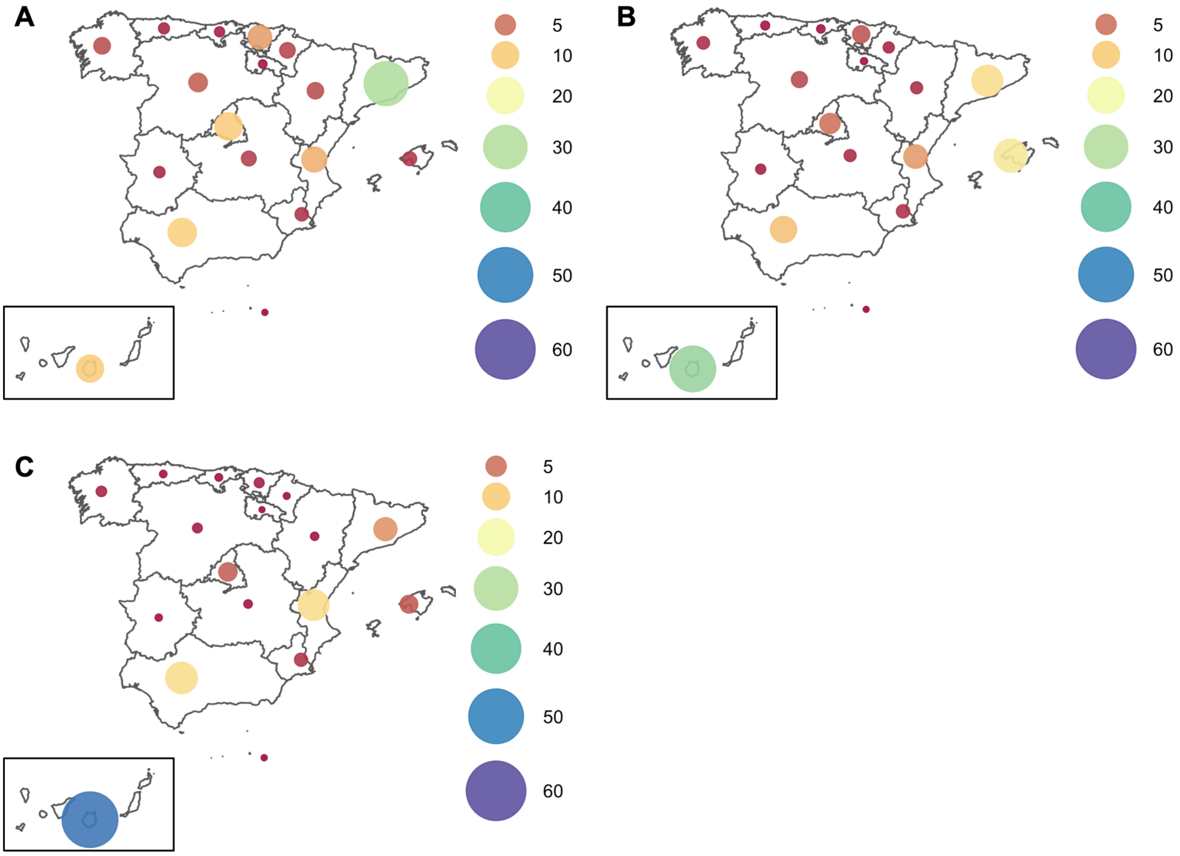


**Figure S5. Connectivity to each Spanish region during the Omicron-BA.1 period** from A) France, B) Germany, and C) the United Kingdom. The colors represent the percentage of total mobility from that country to Spain during this period. The original data were taken from the Spanish National Institute of Statistics (INE; https://www.ine.es).

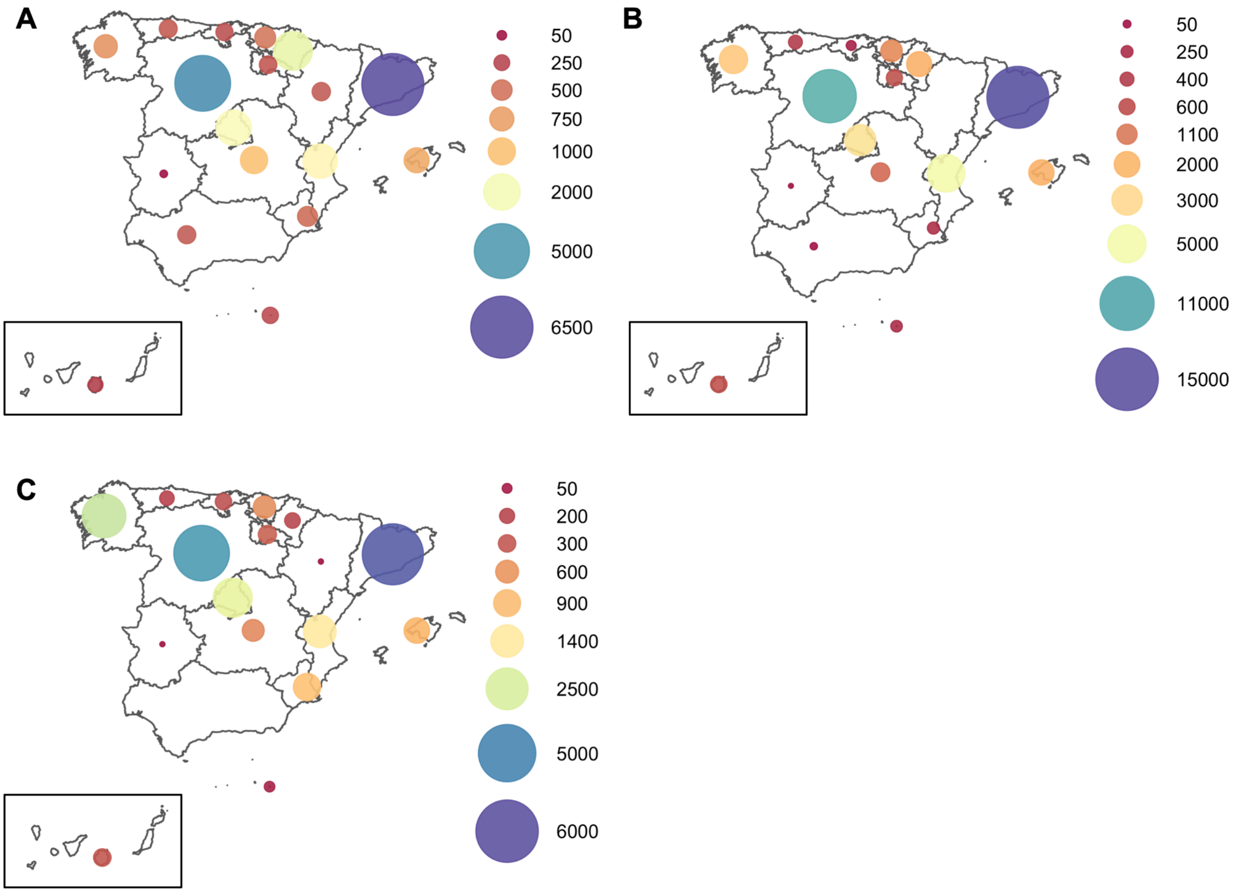


**Figure S6. Number of sequences available in GISAID from each Spanish region, belonging to each of the three variants until 2022-06-11.** A) Alpha. B) Delta. C) Omicron-BA.1. The grey color indicates that no sequences from the specific variant were available at that date in the database.
